# Preliminary modeling estimates of the relative transmissibility and immune escape of the Omicron SARS-CoV-2 variant of concern in South Africa

**DOI:** 10.1101/2022.01.04.22268721

**Authors:** Nicolò Gozzi, Matteo Chinazzi, Jessica T. Davis, Kunpeng Mu, Ana Pastore y Piontti, Alessandro Vespignani, Nicola Perra

**Affiliations:** Networks and Urban Systems Centre, University of Greenwich, UK; Laboratory for the Modeling of Biological and Socio-technical Systems, Northeastern University, Boston, MA USA

**Keywords:** Omicron, B.1.1.529 lineage, SARS-CoV-2, COVID-19, Epidemic modeling

## Abstract

We develop a stochastic, multi-strain, compartmental epidemic model to estimate the relative transmissibility and immune escape of the Omicron variant of concern (VOC) in South Africa. The model integrates population, non-pharmaceutical interventions, vaccines, and epidemiological data and it is calibrated in the period May 1^*st*^, 2021 – November 23^*rd*^, 2021. We explore a parameter space of relative transmissibility with respect to the Delta variant and immune escape for Omicron by assuming an initial seeding, from unknown origin, in the first week of October 2021. We identify a region of the parameter space where combinations of relative transmissibility and immune escape are compatible with the growth of the epidemic wave. We also find that changes in the generation time associated with Omicron infections strongly affect the results concerning its relative transmissibility. The presented results are informed by current knowledge of Omicron and subject to changes.

## Introduction

As of December 28^*th*^, 2021 the Omicron variant of concern (VOC) [1, 2] has been detected in more than 110 countries and territories around the world [3]. The variant was first designated as lineage B.1.1.529 on November 24^*th*^, then named Omicron and classified officially as VOC on November 26^*th*^ [4]. Some of the many mutations carried by the VOC have been linked to escape neutralising antibodies. Several preliminary in vitro experiments confirmed this concerning property of the variant against which even the two dose vaccination appears to be less effective [5, 6]. Vaccines, natural acquired immunity, and boosters however, still offer good protection against severe complications and deaths from the disease [2, 6].

South Africa - in particular the province of Gauteng - was the first country with evidence of Omicron widespread community transmission [2]. At the beginning of May 2021, the country faced a large third wave of infections driven by the Delta VOC. However, since the end of August both cases and deaths have been on a downwards trend [7]. In South Africa only 24% of the population is fully vaccinated as of late November 2021, thus hinting the suppression of the Delta wave, given the lack of very restrictive non-pharmaceutical interventions in the period, was primarily due to high levels of natural immunity across the population. The rapid increase of cases observed in South Africa signalled that the new variant has a marked growth advantage over the Delta VOC. The rapid diffusion of Omicron, in a background of receding Delta, might be explained by increased transmissibility, immune escape, or a combination of both. Early evidence from statistical analysis of possible reinfections, suggests indeed significant capabilities of escape from naturally acquired immunity [7].

Here, we develop a multi-strain, stochastic, compartmental epidemic model for South Africa to identify some of the features of the Omicron VOC compatible with epidemiological observations. The model takes as input data about demographics, age-stratified contact patterns, non-pharmaceutical interventions (NPIs), vaccine rollout, and confirmed COVID-19 deaths and cases (see Materials and Methods for details). We perform a multi-stage calibration applying an Approximated Bayesian Computation (ABC) method [8]. For Omicron, we explore a parameter space defined by the combination of the relative transmissibility with respect to Delta and immune escape with respect to both naturally acquired immunity and vaccines. We obtain the joint posterior distribution of these parameters compatible with the observations for the confirmed cases until December 13^*th*^, 2021 (rescaled to account for under-reporting). The results highlight the non-identifiability of both relative transmissibility and immune escape. Rather, at this stage it is possible to define a region of credible values where an increase in relative transmissibility is compensated by a reduction of immune escape and vice-versa. It is important to notice how the joint posterior distribution is function of the seeding timing and size as well as of the generation time of Omicron infections. Short generation times (i.e., 3.5 days) with respect to the Delta VOC (i.e., 5.5 days) shift the joint posterior distribution to a region with lower values of transmissibility advantage for Omicron. The presented results are informed by current knowledge of the variant and subject to changes as new evidence and data will be available.

## Results

We use a stochastic, multi-strains, and compartmental epidemic model at the national level in South Africa. It features demographics, age-structured contacts patterns, NPIs, vaccines, and epidemiological data on confirmed deaths and cases (see more information in the Material and Methods section). In order to estimate the properties of the Omicron variant we adopt a multi-stage calibration process. Starting around May 2021, the country experienced a third pandemic wave fuelled by the Delta VOC [7]. This variant was able to replace the Beta VOC, which was responsible for the second wave. Before the appearance of Omicron, Delta was responsible for the large majority of cases. Hence, we first fit the model allowing just for one strain in the period May 1^*st*^, 2021 – November 23^*rd*^, 2021. To this end, we adopt the ABC method applied to the weekly confirmed deaths (see Material and Methods). In particular, we obtain the posterior distributions for transmissibility, delay between deaths and their reporting, initial conditions, seasonality, infection fatality rates multiplier respect to the estimates from Ref. [9], and under-reporting in deaths (see Material and Methods for more details). Across the board we estimate that the surveillance system in South Africa was able to detect 1 out of 16 infections in the period from May to November, 2021. These numbers are in line with independent estimates done by Institute for Health Metrics and Evaluation (IHME) which suggests the detection of 1 out of 15 cases [10, 11]. We then study the impact of Omicron by assuming an initial seeding, from unknown origin, in the first week of October 2021. Preliminary evidences from phylogenetic analysis suggest the median date of the common ancestor, of all available Omicron samples, in early October (90% CI: [30 September - 20 October]) [12]. As we do not have information about the number of initial seeds, we sample a flat prior distribution *σ* = [10 − 1000] seeds. We define the Omicron transmissibility by setting 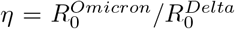 as the ratio of the basic reproductive numbers for Omicron and Delta (see Materials and Methods for more details). The immunity evasion of Omicron is introduced with single factor *α* which describes the reduction of vaccine efficacy and protection from reinfection (see Material and Methods for more details). We explore the ***η*** × ***α*** × ***σ*** parameter space and apply an Approximate Bayesian Computation approach to select values compatible with the evidence for the number of confirmed cases.

### Spreading and immune escape potential

In Figure 1 we plot the joint posterior distribution of *η* and *α* values. We consider two cases: in the first one, Omicron has the same generation time of Delta (*T*_*G*_ = 5.5 days), while in the second one we assume a shorter generation time (*T*_*G*_ = 3.5 days) [13]. We start with a flat three-dimensional prior (on *η, α*, and number of initial seeds). Note how the two-dimensional plot is a projection of the posterior values of the initial seeds. While large regions of the parameter space are rejected by the ABC approach, it emerges a plausible region that corresponds to the non-identifiable nature of both parameters. Generally speaking, the higher the immune escape *α*, the smaller *η*. Intuitively, Omicron’s large value of immune escape can be compensated by changes in the relative transmissibility with respect to Delta and vice versa. The number of initial seeds plays a role. For smaller values of seeds the joint posterior distribution peaks for large value of the relative transmissibility and low immune escape (not shown in the figures). As the number of seeds increases large values of *η* are dropped and the posterior shifts towards a more significant immunity escape. Interestingly, the generation time plays an important role in the values selected by the calibration process. When *T*_*G*_ = 5.5 days, the selected values of *η* go from ∼ 3 when *α* ∼ 0% and approach 1 (i.e., same transmissibility of Delta) for very large values of *α*. For *T*_*G*_ = 3.5 days, the maximum values of relative transmissibility in absence of immune escape are around 2 and fall below 1 (i.e., lower transmissibility of Delta) when *α >* 70%. In the next sections we will consider the case *T*_*G*_ = 5.5 days but very similar conclusions can be drawn in the case of shorter generation time.

**Figure 1:**
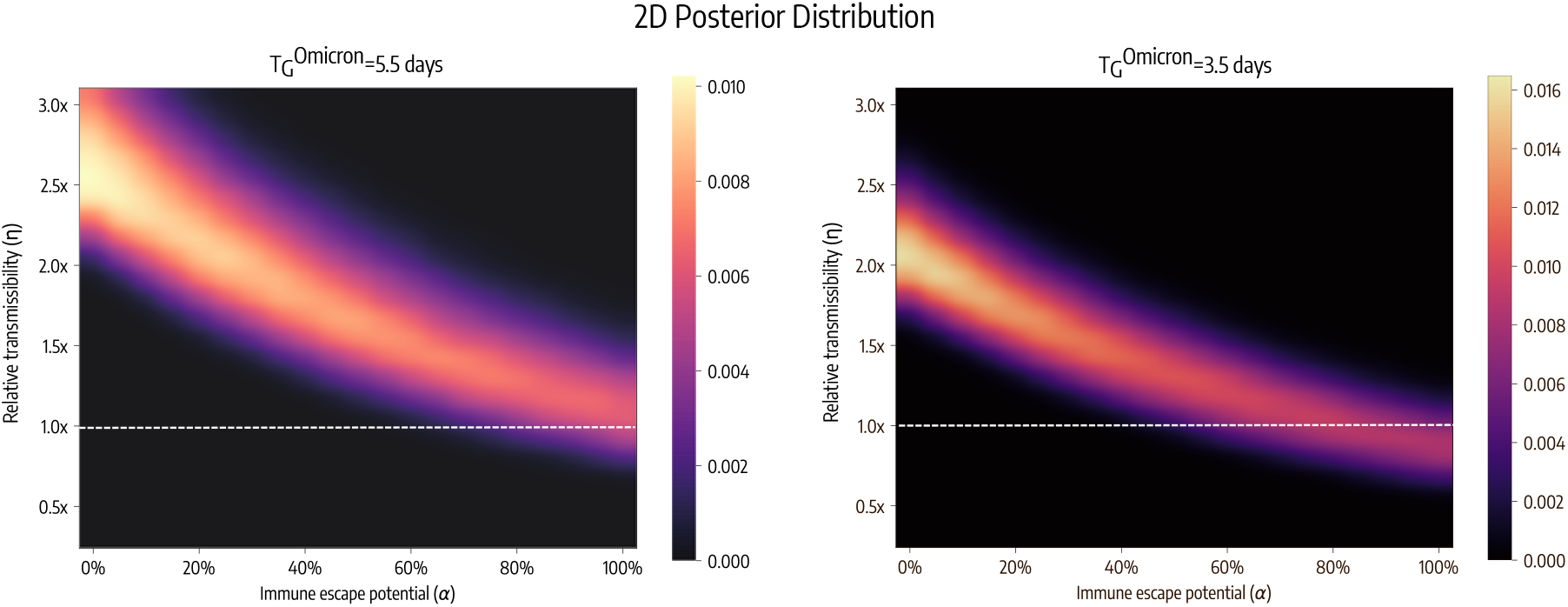
Joint posterior distribution of relative transmissibility (*η*) and immune escape (*α*) for two values of the generation time of Omicron (*T*_*G*_ = 5.5 days on the left and *T*_*G*_ = 3.5 days on the right). The plot highlights the regions of the phase-space compatible with current observations. The colors describe the joint posterior distribution hence the probability that each pair of values is selected in the calibration. The lighter the color the closer the pair of values is to cases. Dashed white horizontal lines indicate the case when Omicron has the same transmission rate as Delta.

### Path towards dominance

In Figure 2 we plot the share of cases due to the Omicron VOC considering the joint posterior distribution for *η, α* and *σ*. The plot suggests that Omicron reached dominance (i.e., more than 50% of cases) in mid-November and replaced Delta by early December. This finding is in line with current genomics data from the sample collected in the region [14]. To put this into perspective, estimates about the time to dominance of the Alpha variant across Europe are between 3 to 4 months [15].

**Figure 2:**
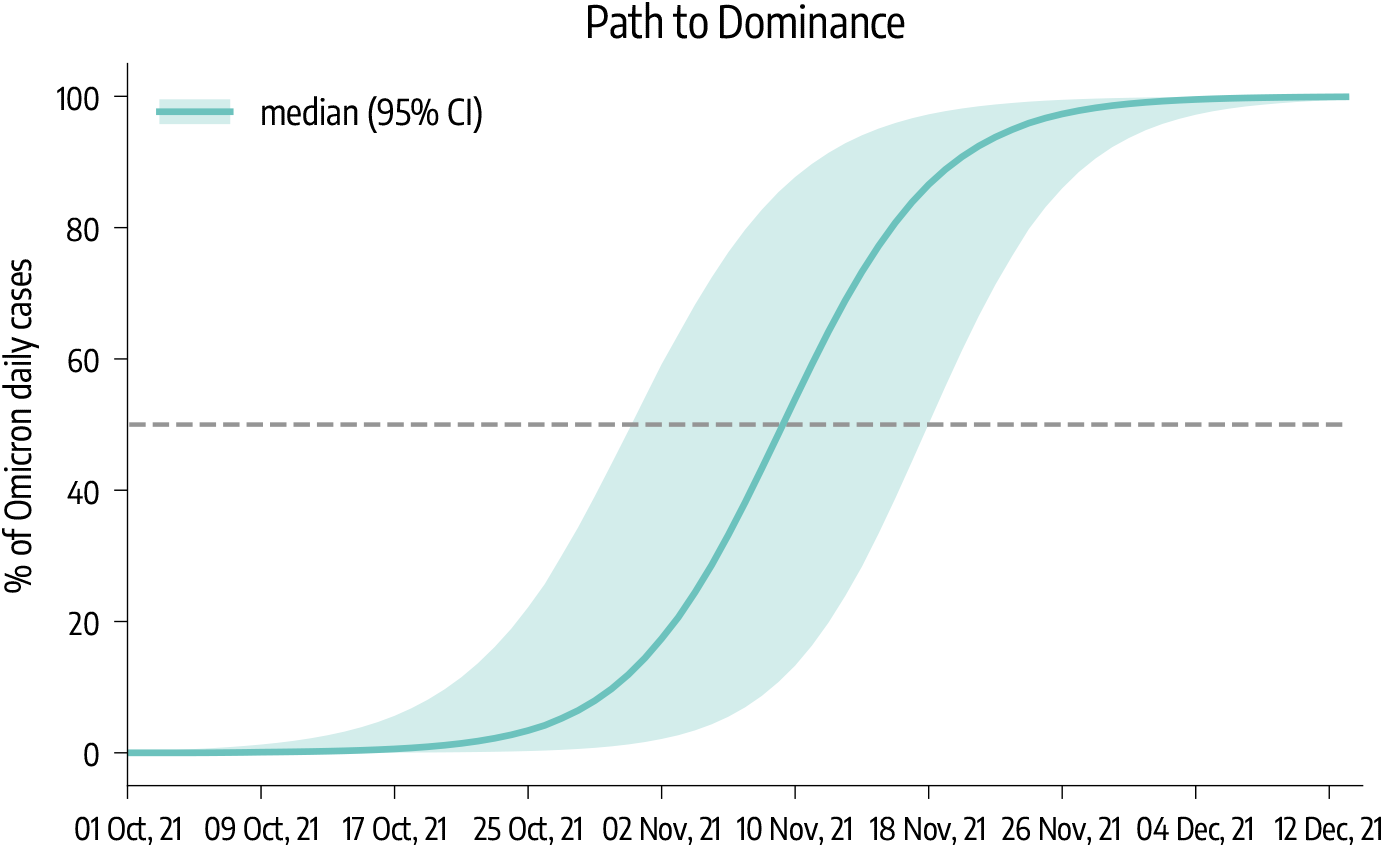
Omicron’s path towards dominance. The plots accounts for all the *η, α* and initial seeds *σ* values selected by the calibration.

### Reproduction number estimates

In Figure 3-A we plot the reproductive numbers *R*_*t*_ (estimated via Epiestim [16]) for the Delta and Omicron VOC assuming the same generation time *T*_*G*_ = 5.5 days. The first observation is that the Delta VOC is subcritical (i.e., *R*_*t*_ ∼ 0.8). This finding is consistent with the decreasing trends before the emergence of Omicron. The values of the reproductive number of Omicron are above 1.6 until the last few data points where we see an inflection. By looking at both variants together we observe how the arrival of Omicron quickly shifted the values of *R*_*t*_ from below one to values around 1.7 in about two months since the seeding. In Figure 3-B, we plot the ratio between the *R*_*t*_ of Omicron and the equivalent quantity for Delta. While we note a rather large confidence intervals the median values are well above 2.2, confirming how the Omicron has a clear growth advantage with respect to Delta in South Africa.

**Figure 3:**
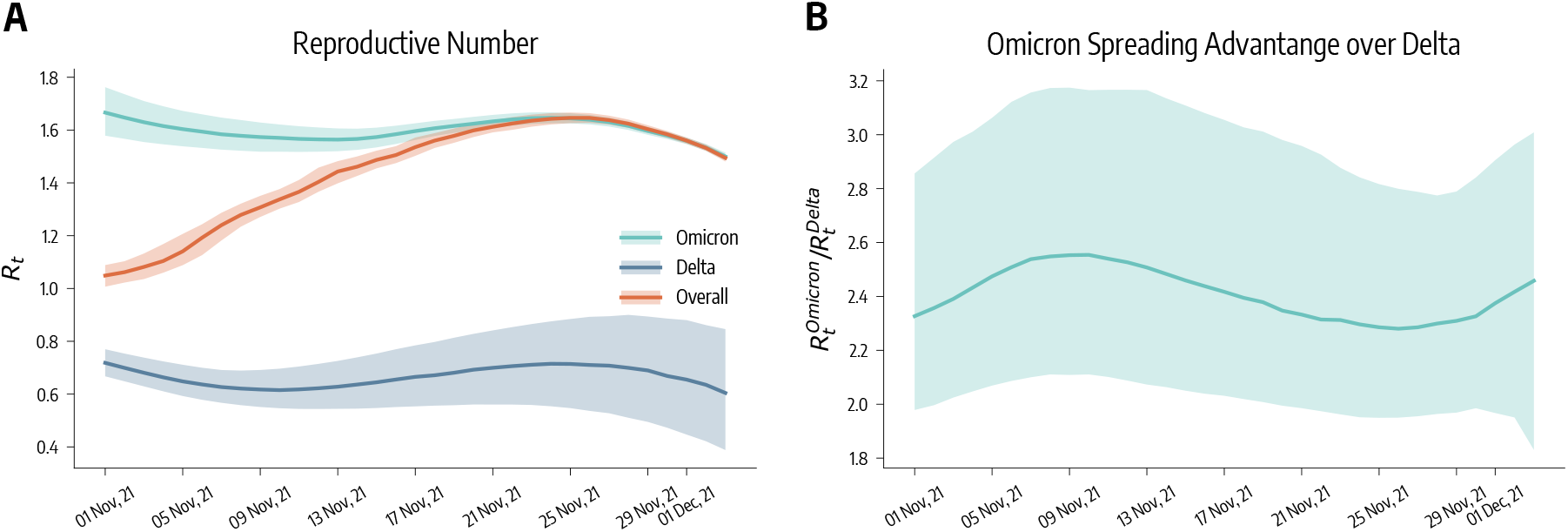
*R*_*t*_ for Omicron, Delta VOC, and for both on the left. Ratio of *R*_*t*_ of the two VOC competing in South Africa. The plots accounts for all the relative transmissibility, immune escape, and initial seeds values selected by the calibration.

## Discussion

Key questions about the Omicron VOC revolve around its relative transmissibility with respect to Delta, the potential for immune escape, and its severity. Although large uncertainties are surrounding Omicron, early statistical analyses suggest the variant is indeed able to reinfect individuals at rates higher than previous VOCs [7] and vaccines might be less effective against infection [5]. Our results are in line with these findings and identify a joint posterior distribution for the relative transmissibility with respect to Delta (*η*), and immune escape (*α*) of the Omicron variant. Current data however does not allow to identify uniquely both parameters, and we find a region where a large spreading advantage might be compensated by a limited immune escape and vice versa. Interestingly, we found that the assumed generation time of Omicron has a significant influence on the results. The findings reinforce the very fast growth of Omicron that is estimated to be the dominant strain in the region since the second week of November (CI [November 2^*nd*^, 2021 – November 17^*th*^, 2021]).

It is important to acknowledge the limitations of our approach. The compartmental structure adopted is relatively simple and does not account explicitly for asymptomatic transmission, and different degrees of severity (i.e., hospitalizations and ICUs). While our model accounts for the COVID-19 vaccine rollout, we have access to limited information about the exact number of different vaccines (i.e., AstraZeneca, J&J, Pfizer, Moderna) administered. For simplicity we assume a two doses regiment across the board.

We set several parameters driving the natural history of the disease using estimates from previous SARS-CoV-2 variants. The large number of mutations and divergence of Omicron could affect some of these values. We model immune escape from previous infections and vaccines with a single parameter. Finally, our model does not consider geographical heterogeneity.

Our results confirm that more data is necessary to estimate the key characteristics of the Omicron VOC. Nevertheless, region of values (capturing its relative transmissibility with respect to Delta and its potential for immune escape) compatible with current observations confirms the likelihood for the Omicron variant of igniting new pandemic waves in regions with high attack rates from previous strains and/or vaccination rates. Data about the severity of Omicron with respect to Delta and the ancestral SARS-CoV-2 viruses are going to be crucial in assessing the impact of on the health-care system of countries affected by surges driven by the Omicorn variant [2, 17].

## Materials and Methods

### Demographic and epidemiological data

We use epidemiological data from the COVID-19 Data Repository by the Center for Systems Science and Engineering (CSSE) at Johns Hopkins University and from official sources [18, 19]. The number of individuals in different age groups is taken from the United Nation World Population Prospects [20].

### Disease transmission model

The disease progression is modeled via a compartmental model. Susceptible and healthy individuals (*S*) interacting with the Infectious (*I*) enter the Latent stage (*L*). After the latent period (*ϵ*^*−*1^), *L* individuals become infectious. Finally, after the infectious period (*µ*^*−*1^), *I* transition to the Recovered (*R*) compartments. Compatibly with the characteristics of the Delta SARS-CoV-2 variant (which was prevalent in South Africa during the period considered before the arrival of Omicron), we set *ϵ*^*−*1^ = 3 days and *µ*^*−*1^ = 2.5 days [21–23]. We consider individuals divided into 10 age groups: [0 − 9, 10 − 19, 20 − 24, 25 − 29, 30 − 39, 40 − 49, 50 − 59, 60 − 69, 70 − 79, 80+], and we describe frequency of contacts between age groups with a country-specific contacts matrix from Ref. [24]. The transmission rate is *β* and the force of infection depends on the age-stratified contact matrix.

We compute the daily number of deaths from individuals entering the *R* compartment using the age-stratified Infection Fatality Rate (IFR) from Ref. [9]. To account for delays due to reporting and hospitalization we record deaths computed on the recovered in day *t* after Δ days. We also introduce a seasonal term to account for changes in factors such as humidity and temperature that can influence transmissibility [25, 26]. This implies a modulation of the effective reproductive number *R*_*t*_ → *s*_*i*_(*t*)*R*_*t*_, with *s*_*i*_(*t*) equal to 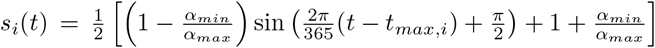, where *i* refers to the hemisphere considered, and *t*_*max,i*_ is the day associated to the maximum of the rescaling function. For the southern hemisphere it is fixed to July 15^*th*^. In the simulations, we set *α*_*max*_ = 1 and consider *α*_*min*_ as a free parameter (see more details below).

We extend this framework with specific compartments to account for vaccinations and the introduction of the Omicron VOC. First, we model vaccinations as follows. Individuals who received the 1^*s*^*t* dose of vaccine transition to a new compartment *V*_1_. Infection probability for *V*_1_ individuals is reduced by a factor 1− *V E*_*S*1_, where *V E*_*S*1_ represents the effectiveness of vaccine against infection. If they get infected their IFR is also reduced by a factor 1 − *V E*_*M*1_. It follows that the overall efficacy of the 1^*st*^ dose against death is *V E*_1_ = 1 − (1 − *V E*_*S*1_)(1 − *V E*_*M*1_). *V*_1_ individuals then receive the second inoculation and transition to the compartment *V*_2_. The 2^*nd*^ dose has an efficacy *V E*_*S*2_ and *V*_*M*2_ (overall efficacy against death *V E*_2_ = 1 − (1 − *V E*_*S*2_)(1 − *V E*_*M*2_)). We consider all vaccinated individuals are less infectious by a factor (1 − *V E*_*I*_) [27]. We assume that *S, L*, and *R* individuals can get the vaccine and since protection is not immediate, we introduce a delay of Δ_*V*_ days between administration (of both 1^*st*^ and 2^*nd*^ dose) and actual effect of the vaccine. We consider the number of 1^*s*^*t* and 2^*nd*^ doses administered daily in South Africa from Ref. [28] and we assume that the vaccine rollout proceeded prioritizing the elderly.

The introduction of a SARS-CoV-2 variant is modeled as follows. We add specific *L* and *I* compartments to account for individuals who are infected with Omicron. We assume that its transmission rate is *ψβ*. For example, *ψ* = 0.5 indicates that the transmission rate of Omicron is half that of Delta, *ψ* = 1.0 the same of Delta, and *ψ* = 2.0 twice that of Delta. In the results presented in Fig. 1 we considered two scenarios with different generation time *T*_*G*_ = 5.5 days, and *T*_*G*_ = 3.5 days. The basic reproductive number in our model is 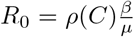, where *ρ*(*C*) is the leading eigenvalue of the contacts matrix. We explore values of the relative transmissibility 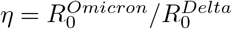 between 0.25 and 3.0 in both scenarios.

In the week November 30^*th*^, 2021 – October 7^*th*^, 2021 we introduce *σ* seeds of the Omicron variant in the infected compartments (half in *I* and half in *L* compartment), and we distribute them in different age groups proportionally to their size. When we introduce the first Omicron infection we also move *R* individuals to a new compartment 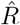. We use a parameter *α* to describe the ability of Omicron VOC to escape natural and vaccine acquired immunity. We assume that individuals in *V*_1_, *V*_2_, and 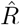 compartments see their protection provided by immunity reduced by a factor (1 − *α*). We assume also that initial protection of 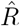 is equal to that of *V*_2_ individuals.

### Modeling of mitigation policies

We quantify the time varying variation in contacts due to mitigation policies by using Google mobility reports [29]. The Google mobility report provides the percentage change *r*_*l*_(*t*) on day *t* of total visitors to specific locations *l* with respect to a pre-pandemic baseline. We turn this quantity into a rescaling factor for contacts such as *ω*(*t*) = (1 + *r*_*l*_(*t*)*/*100)^2^, by considering that the number of potential contacts per location scales as the square of the the number of visitors. The factor *ω*(*t*) is then multiplied to the overall contacts matrix *C*. As *r*_*l*_(*t*) we use the average of the fields `workplaces percent change from baseline, retail and recreation percent change from baseline` and `transit stations percent change from baseline`.

### Model Calibration

We use an Approximate Bayesian Computation (ABC) approach [8, 30]. A prior distribution *P* (*θ*) is defined for the parameters *θ*. At each step of the calibration procedure, a set of parameters 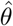 is sampled from *P* (*θ*) an an instance of the model is generated using 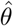. Then an output quantity *E*′ from the model is compared to the corresponding real quantity *E* using a distance metric *s*(*E, E*′). If *s*(*E, E*′) is smaller than a predefined tolerance *δ*, then the sets 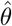 is accepted, otherwise is rejected. The procedure continues until *N* sets are accepted. The empirical distribution of the sampled 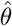 is an approximation of the real posterior distribution of the parameters. Model estimates are then obtained generating an ensemble of trajectories sampled from the approximate posterior. Here we use the weighted mean absolute percentage error (wMAPE) as distance metric and weekly deaths as output quantity, and we set *δ* = 0.25. The parameters *θ* and the related prior distribution are:

- the transmissibility *β*; we explore uniformly values such that the reproductive number on the first day of simulation is between 1.0 and 2.5;
- the seasonality factor *α*_*min*_ ∼ *U* (0.5, 1.0) (0.5 implies strong seasonality while 1.0 no seasonality);
- the delay in deaths Δ ∼ *U* (10, 30) (discrete values);
- the initial number of infected individuals; we explore uniformly values between 1 and 10 times the number of reported cases in the week before the start of the simulation. We then assign these individuals to the infected compartments (*L, I*) proportionally to the time spent there by individuals (*ϵ*^*−*1^ for *L* and *µ*^*−*1^ for *I*);
- the initial number of recovered; we explore uniformly values between 2 and 20 times the total number of reported cases up to the start of the simulation;
- the percentage of deaths that are reported ∼ *U* (25%, 100%);
- a multiplier of the IFR ∼ *U* (0.5, 2.0)

The model is calibrated over the period May 1^*st*^, 2021 – November 23^*rd*^, 2021.

We use the ABC approach also to identify the posterior of Omicron parameters: relative transmissibility respect to Delta 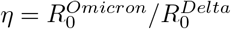, immune escape potential *α*, and number of initial seeds *σ*. We consider values of *η* between 0.25 and 3.0, values of *α* between 0% and 100%, and 10, 50, 100, 500, 1000 initial Omicron seeds introduced in first week of October 2021. We repeat the simulations over this grid of (*η, α, σ*) values during the period November 26^*th*^, 2021 – December 13^*th*^, 2021 and we compare the simulated incidence to the reported daily incidence in South Africa using the wMAPE. For each triplet (*η, α, σ*), we perform 1500 stochastic realizations and we compute the posterior distribution considering the top 5% runs.

## Data Availability

All data produced in the present study are available upon request to the authors

## Acknowledgements

All authors thank the High Performance Computing facilities at Greenwich University. N.G. acknowledges support from the Doctoral Training Alliance. MC, and AV acknowledge support from COVID Supplement CDC-HHS-6U01IP001137-01. MC and AV acknowledge support from Google Cloud and Google Cloud Research Credits program to fund this project. The findings and conclusions in this study are those of the authors and do not necessarily represent the official position of the funding agencies, the National Institutes of Health, or the U.S. Department of Health and Human Services.

## References

[1] Ewen Callaway. Heavily mutated coronavirus variant puts scientists on alert. Nature.

[2] Implications of the further emergence and spread of the SARS-CoV-2 B.1.1.529 variant of concern (Omicron) for the EU/EEA – first update. https://www.ecdc.europa.eu/sites/default/files/documents/threat-assessment-covid-19-emergence-sars-cov-2-variant-omicron-december-2021.pdf, 2021. Accessed: 2021-12-02.

[3] Tracking COVID-19 variant Omicron. https://newsnodes.com/omicrontracker/, 2021.

[4] World Health Organization (WHO). Classification of Omicron (B.1.1.529): SARS-CoV-2 Variant of Concern. https://www.who.int/news/item/26-11-2021-classification-of-omicron-(b.1.1.529)-sars-cov-2-variant-of-concern, 2021. Accessed: 2021-12-02.

[5] Sivan Gazit, Roei Shlezinger, Galit Perez, Roni Lotan, Asaf Peretz, Amir Ben-Tov, Dani Cohen, Khitam Muhsen, Gabriel Chodick, and Tal Patalon. Comparing sars-cov-2 natural immunity to vaccine-induced immunity: reinfections versus breakthrough infections. medRxiv, 2021.

[6] Ewen Callaway. Tracking COVID-19 variant Omicron. Nature, 2021.

[7] Juliet R.C. Pulliam, Cari van Schalkwyk, Nevashan Govender, Anne von Gottberg, Cheryl Cohen, Michelle J. Groome, Jonathan Dushoff, Koleka Mlisana, and Harry Moultrie. Increased risk of sars-cov-2 reinfection associated with emergence of the omicron variant in south africa. medRxiv, 2021.

[8] Mikael Sunnåker, Alberto Giovanni Busetto, Elina Numminen, Jukka Corander, Matthieu Foll, and Christophe Dessimoz. Approximate bayesian computation. PLOS Computational Biology, 9(1):1–10, 01 2013.

[9] Robert Verity, Lucy Okell, Ilaria Dorigatti, Peter Winskill, Charles Whittaker, Natsuko Imai, Gina Cuomo-Dannenburg, Hayley Thompson, Patrick Walker, Han Fu, Amy Dighe, Jamie Griffin, Marc Baguelin, Sangeeta Bhatia, Adhiratha Boonyasiri, Anne Cori, Zulma M. Cucunubá, Rich FitzJohn, Katy Gaythorpe, and Neil Ferguson. Estimates of the severity of coronavirus disease 2019: a model–based analysis. The Lancet Infectious Diseases, 20, 03 2020.

[10] How epidemiological models of COVID-19 help us estimate the true number of infections. https://ourworldindata.org/covid-models, 2021. Accessed: 2021-12-14.

[11] COVID-19 Projections, South Africa. https://covid19.healthdata.org/south-africa?view=cumulative-deaths&tab=trend, 2021. Accessed: 2021-12-14.

[12] Rapid epidemic expansion of the SARS-CoV-2 Omicron variant in southern Africa. https://ceri.africa/publication/?token=369, 2021. Accessed: 2021-12-14.

[13] Lin T. Brandal, Emily MacDonald, Lamprini Veneti, Tine Ravlo, Heidi Lange, Umaer Naseer, Siri Feruglio, Karoline Bragstad, Olav Hungnes, Liz E. Ødeskaug, Frode Hagen, Kristian E. Hanch-Hansen, Andreas Lind, Sara Viksmoen Watle, Arne M. Taxt, Mia Johansen, Line Vold, Preben Aavitsland, Karin Nygård, and Elisabeth H. Madslien. Outbreak caused by the sars-cov-2 omicron variant in norway, november to december 2021. Eurosurveillance, 26(50), 2021.

[14] Variant: 21K (Omicron). https://covariants.org/variants/21K.Omicron, 2021.

[15] Nicolò Gozzi, Matteo Chinazzi, Jessica T Davis, Kunpeng Mu, Ana Pastore y Piontti, Marco Ajelli, Nicola Perra, and Alessandro Vespignani. Estimating the spreading and dominance of sars-cov-2 voc 202012/01 (lineage b. 1.1. 7) across europe. medRxiv, 2021.

[16] Anne Cori, Neil M. Ferguson, Christophe Fraser, and Simon Cauchemez. A New Framework and Software to Estimate Time-Varying Reproduction Numbers During Epidemics. American Journal of Epidemiology, 178(9):1505–1512, 09 2013.

[17] Neil ferguson, azra ghani, wes hinsley and erik volz. hospitalisation risk for omicron cases in england. https://www.imperial.ac.uk/media/imperial-college/medicine/mrc-gida/2021-12-22-COVID19-Report-50.pdf, 2021. Accessed: 2021-12-23.

[18] COVID-19 Data Repository by the Center for Systems Science and Engineering (CSSE) at Johns Hopkins University. https://github.com/CSSEGISandData/COVID-19, 2021.

[19] COVID-19 Online Resources, South Africa. https://sacoronavirus.co.za, 2021.

[20] united nations, department of economic and social affairs, population division. world population prospects: The 2019 revision.

[21] Jantien A Backer, Don Klinkenberg, and Jacco Wallinga. Incubation period of 2019 novel coronavirus (2019-ncov) infections among travellers from wuhan, china, 20–28 january 2020. Eurosurveillance, 25(5), 2020.

[22] Stephen M. Kissler, Christine Tedijanto, Edward Goldstein, Yonatan H. Grad, and Marc Lipsitch. Projecting the transmission dynamics of sars-cov-2 through the postpandemic period. Science, 368(6493):860–868, 2020.

[23] Baisheng Li, Aiping Deng, Kuibiao Li, Yao Hu, Zhencui Li, Qianling Xiong, Zhe Liu, Qianfang Guo, Lirong Zou, Huan Zhang, Meng Zhang, Fangzhu Ouyang, Juan Su, Wenzhe Su, Jing Xu, Huifang Lin, Jing Sun, Jinju Peng, Huiming Jiang, Pingping Zhou, Ting Hu, Min Luo, Yingtao Zhang, Huanying Zheng, Jianpeng Xiao, Tao Liu, Rongfei Che, Hanri Zeng, Zhonghua Zheng, Yushi Huang, Jianxiang Yu, Lina Yi, Jie Wu, Jingdiao Chen, Haojie Zhong, Xiaoling Deng, Min Kang, Oliver G. Pybus, Matthew Hall, Katrina A. Lythgoe, Yan Li, Jun Yuan, Jianfeng He, and Jing Lu. Viral infection and transmission in a large, well-traced outbreak caused by the sars-cov-2 delta variant. medRxiv, 2021.

[24] Dina Mistry, Maria Litvinova, Ana Pastore y Piontti, Matteo Chinazzi, Laura Fumanelli, Marcelo F C Gomes, Syed A Haque, Quan-Hui Liu, Kunpeng Mu, Xinyue Xiong, M Elizabeth Halloran, Ira M Longini, Stefano Merler, Marco Ajelli, and Alessandro Vespignani. Inferring high-resolution human mixing patterns for disease modeling. Nature Communications, 12(1):323, 2021.

[25] Duygu Balcan, Bruno Gonçalves, Hao Hu, José J. Ramasco, Vittoria Colizza, and Alessandro Vespignani. Modeling the spatial spread of infectious diseases: The global epidemic and mobility computational model. Journal of Computational Science, 1(3):132 – 145, 2010.

[26] Ben S Cooper, Richard J Pitman, W John Edmunds, and Nigel J Gay. Delaying the international spread of pandemic influenza. PLoS Med, 3(6):e212, 2006.

[27] Julia Shapiro, Natalie E. Dean, Zachary J. Madewell, Yang Yang, M.Elizabeth Halloran, and Ira Longini. Efficacy estimates for various covid-19 vaccines: What we know from the literature and reports. medRxiv, 2021.

[28] Edouard Mathieu, Hannah Ritchie, Esteban Ortiz-Ospina, Max Roser, Joe Hasell, Cameron Appel, Charlie Giattino, and Lucas Rodés-Guirao. A global database of COVID-19 vaccinations. Nature Human Behaviour, 5(7):947–953, 2021.

[29] Google LLC “Google COVID-19 Community Mobility Reports”. https://www.google.com/covid19/mobility/, 2020. Accessed: 2021-08-01.

[30] Amanda Minter and Renata Retkute. Approximate bayesian computation for infectious disease modelling. Epidemics, 29:100368, 2019.

